# Determinants of losses in the latent tuberculosis infection cascade of care in Brazil: a prospective multicenter cohort study

**DOI:** 10.1101/2021.03.12.21252212

**Authors:** Alexandra Brito Souza, María B. Arriaga, Gustavo Amorim, Mariana Araújo-Pereira, Betânia M. F. Nogueira, Artur T. L. Queiroz, Marina C. Figueiredo, Michael S. Rocha, Aline Benjamin, Adriana S. R. Moreira, Jamile G. de Oliveira, Valeria C. Rolla, Betina Durovni, José R. Lapa e Silva, Afrânio L. Kritski, Solange Cavalcante, Timothy R. Sterling, Bruno B. Andrade, Marcelo Cordeiro-Santos, for the RePORT Brazil consortium

## Abstract

**Background:** Factors associated with losses in the latent tuberculosis infection (LTBI) cascade of care in contacts of tuberculosis (TB) patients were investigated in a multicenter prospective cohort from highly endemic regions in Brazil.

**Methods:** Close contacts of 1,187 culture-confirmed pulmonary TB patients were prospectively studied between 2015 and 2019, with follow-up between 6 and 24 months. Data on TB screening by clinical investigation, radiographic examination and interferon-gamma release assay (IGRA) were collected. Stepwise multivariable models were used to identify determinants of losses in the LTBI cascade.

**Results:** Among 4,145 TB contacts initially identified, 1,901 were examined (54% loss). Within those individuals, 933 were people living with HIV, ≤5 years-old and/or had positive IGRA results, therefore having recommendation to start TB preventive treatment (TPT). Of those, 454 (23%) initiated treatment, and 247 (6% of all TB contacts) completed TPT. Multivariable regression analysis revealed that living with HIV, illiteracy, and black/*pardo* (brown) race were independently associated with losses in cascade.

**Conclusion:** There were losses at all LTBI cascade stages, but particularly at the initial screening and examination steps. Close contacts who are socially vulnerable and living with HIV were at heightened risk of not completing the LTBI cascade of care in Brazil.

**40-word summary of the article’s main point:** We investigated factors associated with losses in the latent tuberculosis infection cascade of care in a large cohort of contacts in Brazil. Social vulnerability and HIV infection were the most relevant determinants of losses in the LTBI cascade of care.

## INTRODUCTION

The United Nations (UN) and the World Health Organization (WHO) have set ambitious targets for reducing the global burden of TB by 2030 and recognize the essential role of diagnosis and treatment of latent TB infection (LTBI) as a strategy for TB control and elimination [1, 2]. TB preventive therapy (TPT) entails treatment of LTBI to prevent progression to active disease [1].

The impact of TPT in reducing TB incidence at a community level varies according to the epidemiological scenario. It can be a very effective strategy in low burden countries, where the risk of reinfection is low. In the US, for instance, nearly 85% of TB cases originate from reactivation of LTBI [3], and a significant reduction in the number of cases may be achieved with expansion of testing and treatment of LTBI, particularly in the foreign-born population, which accounts for 70% of TB cases [4, 5]. In Brazil, a cluster-randomized trial of LTBI diagnosis and treatment in HIV clinics led to a significant reduction in TB incidence and death [6].

Recent studies conducted in high-income and low- and middle-income countries indicate that important losses occur at all stages of the LTBI cascade for care [7]. A study conducted in 12 health facilities in three Brazilian cities with high TB incidence rates found that most losses in the cascade occurred in the first two steps, which were contact identification and tuberculin skin testing (TST) [8]. Another study found that few people who had a positive TST were started on TPT, and the completion rate was also low [9]. In a study carried out in a pediatric hospital in the state of Rio de Janeiro, Brazil, there was an association between low human development index and loss to follow-up during TPT in children and adolescents [10].

Brazil is one of the 30 high TB-burden nations, and has the largest number of cases in the Americas [2]. Despite a strong National TB Control Program, universal free healthcare for all citizens, and a reliable supply of anti-TB drugs, rates of TB have changed little in recent years [11]. The Brazilian Ministry of Health recommends TPT for all TB contacts with a positive interferon-gamma release assay (IGRA) or TST, as well as to all close contacts who are children ≤ 5 years old or people living with HIV (PLWH), regardless of the test results [11]. The LTBI care cascade in Brazil and the factors associated with losses are not fully understood, as most of the studies to date have been retrospective or cross-sectional. The current study was conceptualized to fill this important gap in the knowledge, to help the design decision-making strategies to improve the TB control. To do so, we aimed to evaluate the LTBI care cascade in a prospective Brazilian cohort of contacts of patients with culture-confirmed pulmonary TB, and identify factors associated with losses at each phase of the cascade.

## METHODS

### Ethics Statement

The protocol, informed consent, and study documents were approved by the institutional review boards at all study sites. The Institional Review Boards and the protocol approval numbers are as follows: (i) Instituto de Pesquisa Clínica Evandro Chagas, Fundação Oswaldo Cruz, Rio de Janeiro, Brazil (protocol no. 688.067), (ii) Secretaria Municipal de Saude do Rio de Janeiro, Brazil (protocol no. 740.554), (iii) Hospital Universitario Clementino Fraga Filho Rio de Janeiro, Brazil (protocol no. 852.519), (iv) Maternidade Climério de Oliveira, Universidade Federal da Bahia, Salvador, Brazil (protocol no. 723.168), (v) Fundação de Medicina Tropical Dr. Heitor Vieira Dourado, Manaus, Brazil (protocol no. 807.595). Participation was voluntary and written informed consent was obtained from all participants.

### Study design

In this study, we included data from close TB contacts identified in the Regional Prospective Observational Research in Tuberculosis (RePORT)-Brazil [12] between August 27^th^, 2015, and July 18^th^, 2019, with follow-up between 6 and 24 months. The RePORT-Brazil consortium is an ongoing, multicenter, cohort study, which follows culture-confirmed pulmonary TB cases and their close contacts. Enrollment sites include The RePORT-Brazil activities take place in three Brazilian states, recruiting participants from a total of five health units: Instituto Nacional de Infectologia Evandro Chagas, Clínica da Familia Rinaldo Delamare, and Secretaria de Saúde de Duque de Caxias (all from Rio de Janeiro state), Instituto Brasileiro para Investigação da Tuberculose (Bahia), and Fundação Medicina Tropical Dr. Heitor Vieira Dourado (Amazonas).

For the present study, contacts were eligible to participate if exposed to an index case of culture-confirmed pulmonary TB who enrolled into RePORT-Brazil and had no evidence of active TB. Exposure to the index TB case was defined as at least 4 hours in one week in the 6 months prior to TB diagnosis.

After enrollment of the index case, close contacts were invited to be interviewed and examined at the RePORT-Brazil healthcare units by phone call, text message or in person. Eligible contacts who attended the study sites were approached by study personnel to enroll into the RePORT-Brazil cohort and to be investigated for LTBI. The contact investigation included clinical evaluation, chest X-ray, blood collection for IGRA and HIV testing. IGRA collection, processing and interpretation were performed according to the manufacturer’s recommendations for QuantiFERON^®^ assay (Qiagen). Clinical and demographic data were collected via standardized case report forms. Per the RePORT-Brazil protocol, all contacts with a negative or indeterminate baseline IGRA test underwent repeat testing at month 6.

TPT was recommended according to the National TB Control Program of Brazil [2, 11]. TPT was also initiated based on the physician’s decision to treat and RePORT-Brazil did not influence such decision.

### Outcome definition

For this analysis the following steps of the LTBI cascade of care were considered: i) identified by TB patients as close contacts, to be screened for LTBI; ii) presented to clinic and agreed to participate, signed informed consent, and had IGRA performed; iii) completed medical and radiographic evaluations; iv) recommended to receive TPT; v) accepted and initiated TPT; vi) completed TPT (defined as: ≤ 6 months of isoniazid or 4 months of rifampicin).

### Data analysis

Categorical variables were presented as frequencies and compared using Fisher’s exact test (2×2) or Pearson’s chi-square test. Continuous variables were described using median and interquartile ranges and compared using the Mann-Whitney *U* test (between two groups) or the Kruskal Wallis test (two or more groups).

A multivariable generalized estimating equation [13] with a logit link and an independent working correlation matrix was used to identify factors associated with losses in the LTBI cascade of care. Close contacts from the same TB index case were treated as a cluster. Multiple imputation by chained equations [14] was used to generate 20 imputed datasets and final estimates were obtained via Rubin’s rule [15]. Hierarchical cluster analysis (Ward’s method) was employed to depict the overall profile of the study subgroups stratified according to the IGRA final status.

Data analysis was performed using SPSS (IBM Corp. Released in 2015. IBM SPSS Statistics for Windows, Version 25.0, GraphPad Prism 8.0 (GraphPad Software, La Jolla, CA, USA), JMP Pro 14.0, and R 3.5.0 (R Foundation, Vienna, Austria). All analyses were pre-specified and two-tailed. Differences with p-value <0.05 were considered statistically significant.

## RESULTS

### Characteristics of the study participants

There were 4,145 close contacts referred from 1,187 pulmonary TB (PTB) cases (average of 3.5 contacts for each TB case), but only 2,483 contacts of 641 culture confirmed PTB cases could be reached for invitation to present at the health care units and showed up for LTBI screening (**Figure 1**). Of the 2,483 contacts, 1901 (74%) provided informed consent, and completed the medical evaluation. The characteristics of TB contacts are presented in **Table 1**. Median age was 32 years (IQR 16.2-46.8), 1,130 (59%) were female, 1507 (79%) were black/*pardo* (brown), 192 (10%) were illiterate and 46 (2%) were PLWH. A positive IGRA result was detected in 43% of the study participants, including results from baseline and month 6. Participants with a positive IGRA were more likely to report smoking (30% vs. 24%; p=0.002), and secondary smoking (35% vs. 30%; p=0.04) than persons with a negative IGRA result.

**Table 1.** Characteristics of the TB contacts by IGRA result

| Characteristics | Negative<br>(n=1059, 57%) | Positive<br>(n=813, 43%) | p-value |
| --- | --- | --- | --- |
| **Female– no. (%)** | 606 (57) | 509 (63) | 0.002 |
| **Age – median (IQR)** | 30 (15-44) | 36 (18-50) | 0.004 |
| **Race/Ethnicity – no. (%)** |  |  | 0.003 |
| Black/ Pardo | 813 (77) | 671 (83) |  |
| Others<sup>a</sup> | 245 (23) | 142 (17) |  |
| **Income – no. (%)** |  |  | 0.954 |
| More than a minimum wage | 409 (40) | 276 (35) |  |
| Equal or less than a minimum wage | 388 (38) | 357 (45) |  |
| Without income | 225 (22) | 155 (20) |  |
| **BCG scar – no. (%)** | 950 (90) | 721 (89) | 0.498 |
| **HIV infection – no. (%)** | 32 (3) | 14 (2) | 0.097 |
| **Antiretroviral therapy – no. (%)** | 26 (8) | 10 (7) | 0.465 |
| **Education– no. (%)** |  |  | 0.137 |
| Literate | 943 (89) | 743 (91) |  |
| Illiterate | 114 (11) | 70 (9) |  |
| **Smoking – no. (%)** | 253 (24) | 246 (30) | 0.002 |
| **Secondary smoking – no. (%)** | 319 (30) | 281 (35) | 0.04 |
| **Alcohol consumption – no. (%)** | 565 (53) | 442 (54) | 0.674 |
| **Alcohol consumption (years)-median (IQR)** | 10 (3-18) | 14 (5-25) | 0.002 |
| **CAGE score of 2 and above<sup>b</sup> – no. (%)** | 110 (30) | 67 (24) | 0.075 |
| **Illicit drug use – no. (%)** | 102 (10) | 84 (10) | 0.640 |
| **Comorbidities<sup>c</sup>– no. (%)** | 242 (23) | 199 (245) | 0.411 |
| Diabetes | 58 (6) | 34 (4) | 0.235 |
| Hypertension | 120 (11) | 111 (14) | 0.137 |

**Table note:** Data represent no. (%). or median with Interquartile range (IQR)
<sup>a</sup> Others ethnicities: White, Asian and Indian.
<sup>b</sup> Alcohol abuse was defined as a CAGE score $\geq 2$ points as described in Methods. Continuous variables were compared using the Mann-Whitney *U* test and categorical variables were using the Fisher's exact test (2x2) or Pearson's chi-square test.
Missing information was identified in 3 variables: Race (1 [0.05%]), Income (62 [ 3.3%]) and education (2 [0.1%]). The statistical analyzes were carried out only with the available data, omitting the cases with missing information.
<sup>c</sup> Comorbidities: At least one comorbidity (diabetes, hypertension, cancer, chronic obstructive pulmonary /emphysema, kidney disease, heart disease, liver disease and depression).
\*29 (1.5%) individuals had an indeterminate result in the 1st or 2nd IGRA.

**Figure 1.**
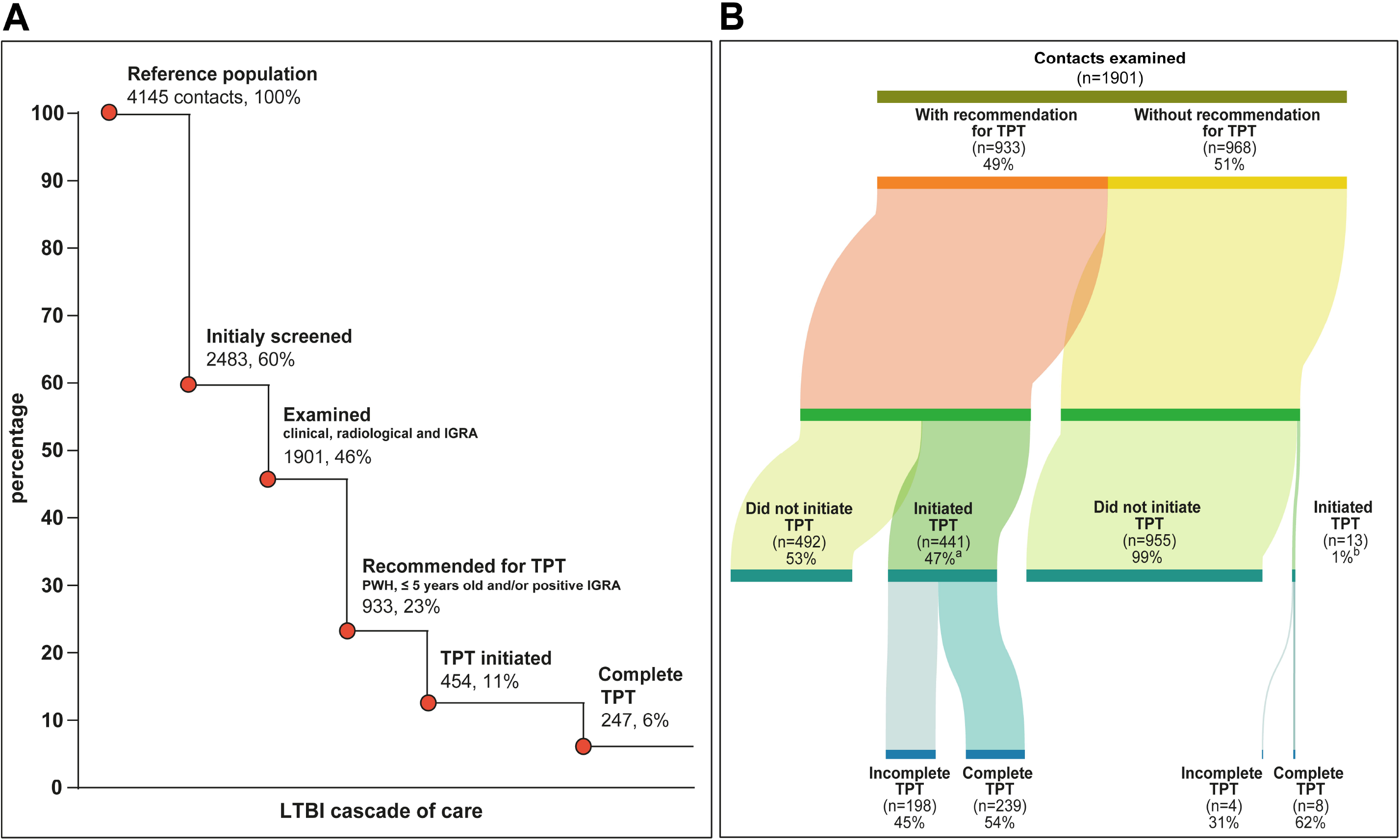
Cascade of care of Latent Tuberculosis Infection in contacts from TB cases. **(A)** Losses and drop-outs at each stage of the LTBI cascade of care. Percentages were calculated among the number of contacts initially identified (n = 4145). **(B)** This figure shows the number of contacts who started treatment and those who completed treatment according to the category of TPT recommendation. ^a^4 contacts were still undergoing TPT at the time of analysis (not considered losses). ^b^1 contact was still undergoing TPT at the time of analysis (not considered loss). Abbreviations: IGRA: Interferon-gamma release assay; PWH: People with HIV; LTBI: Latent tuberculosis infection. TB: tuberculosis. TPT: TB preventive therapy.

### Losses of LTBI Cascade of care

The losses in the LTBI cascade are presented in **Figure 1**. We observed losses in all steps of the cascade. Most losses occurred early in the cascade, where 40% of the identified contacts were not evaluated for screening, and another 14% were not examined (**Figure 1A**). Of the 933 contacts who met criteria for receiving TPT, 441 (47%) initiated TPT (**Figure 1B**). Of the 441 who initiated treatment, 239 (54% of those initiating; 26% of those eligible; 6% of all contacts) completed it. Among all study participants, factors associated with overall losses in the LTBI cascade were black/*pardo* race (p=0.024), a wage ≤ the minimum established for Brazil (p=0.009), HIV infection (p=0.018) and to be illiterate (p<0.001) (**Supplementary Table 1**).

Analyses of the time from detection of PTB index cases to screening of contacts are presented in (**Figure 2)**. Contacts were evaluated on average approximately 1 month after the diagnosis of the TB index case (median=36 days) (**Figure 2A and 2B**). Of note, contacts with different outcomes in the LTBI cascade could not be distinguished based on time from identification of the TB index case and the first screening for LTBI (**Figure 2B and 2C**).

**Figure 2.**
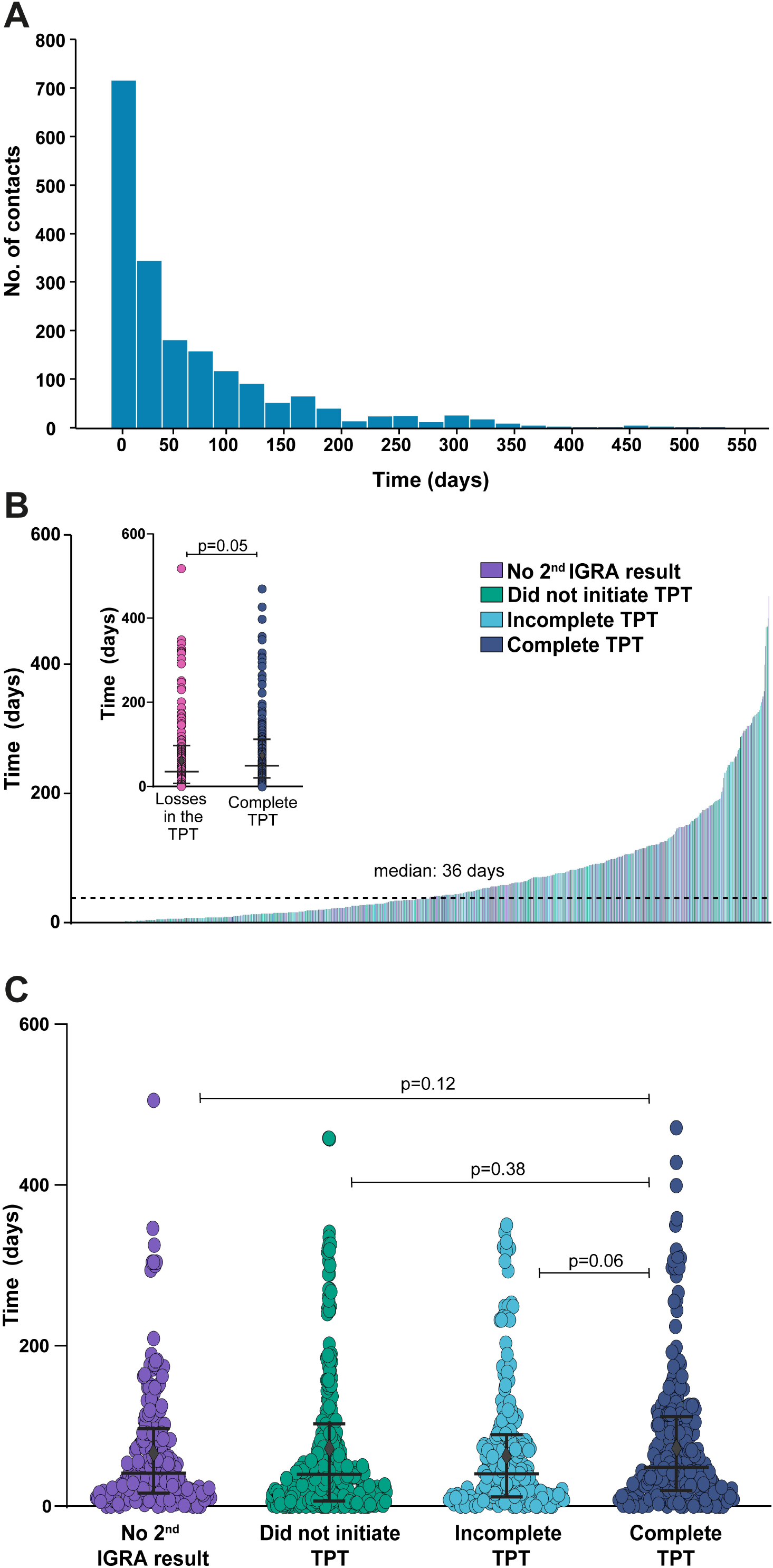
Time from detection of TB index cases to screening of their contacts. **(A)** Histograms representing the frequency of TB contacts between time (days) from visit 1 of the TB index case and visit 1 of the contact. (B) Histograms representing time (days) between visit 1 of the TB index case and visit 1 of the contact by type of losses in the LTBI cascade for care. **(C)** Scatter plots show each contact and case and the time difference between visit 1 of the TB index case and visit 1 of the contact by type of losses in the LTBI cascade for care. Did not perform 2nd IGRA (n=236), did not initiate recommended TPT (n=493), did not complete TPT (n=202) and complete TPT (n=247). Abbreviations: IGRA: interferon-gamma release assay. LTBI: Latent tuberculosis infection. TB: tuberculosis. TPT: TB preventive therapy.

### Factors associated with overall losses of LTBI cascade of care

**Figure 3** presents associations between clinical and demographic factors and overall losses in the LTBI cascade for care (i.e., all losses in the LTBI cascade combined). There were no significant differences in sex distribution or in the ages of patients who were lost in the LTBI cascade or completed treatment. However, in univariate analyses, race black or pardo (p=0.024), low personal income (p=0.009) and illiteracy (p<0.001) were associated with losses in the LTBI cascade of care **(Supplementary Table 1, Figures 3B and 3C**). Multivariable analyses also revealed that illiteracy (adjusted Odds Ratio [aOR]: 3.67, 95% Confidence Interval [CI]: 2.40-5.62, p > 0.001), HIV infection (aOR: 2.63, 95% CI: 1.08-6.4, = 0.032) and race black or pardo (aOR: 1.33, 95% CI: 1.06-1.67, p = 0.014) were independently associated with losses in the LTBI cascade (**Figure 3C**). The impact of time from the identification of the TB index case to the initial screening of the close contact on the overall losses in the LTBI cascade was only marginal, but statistically significant (**Figure 3C**). Indeed, each increase in one day between the PTB diagnosis and the enrollment of the close contact resulted in an adjusted odds of 0.99 (95% CI: 0.98-1.0, p=0.032) of being lost in the LTBI cascade of care.

**Figure 3.**
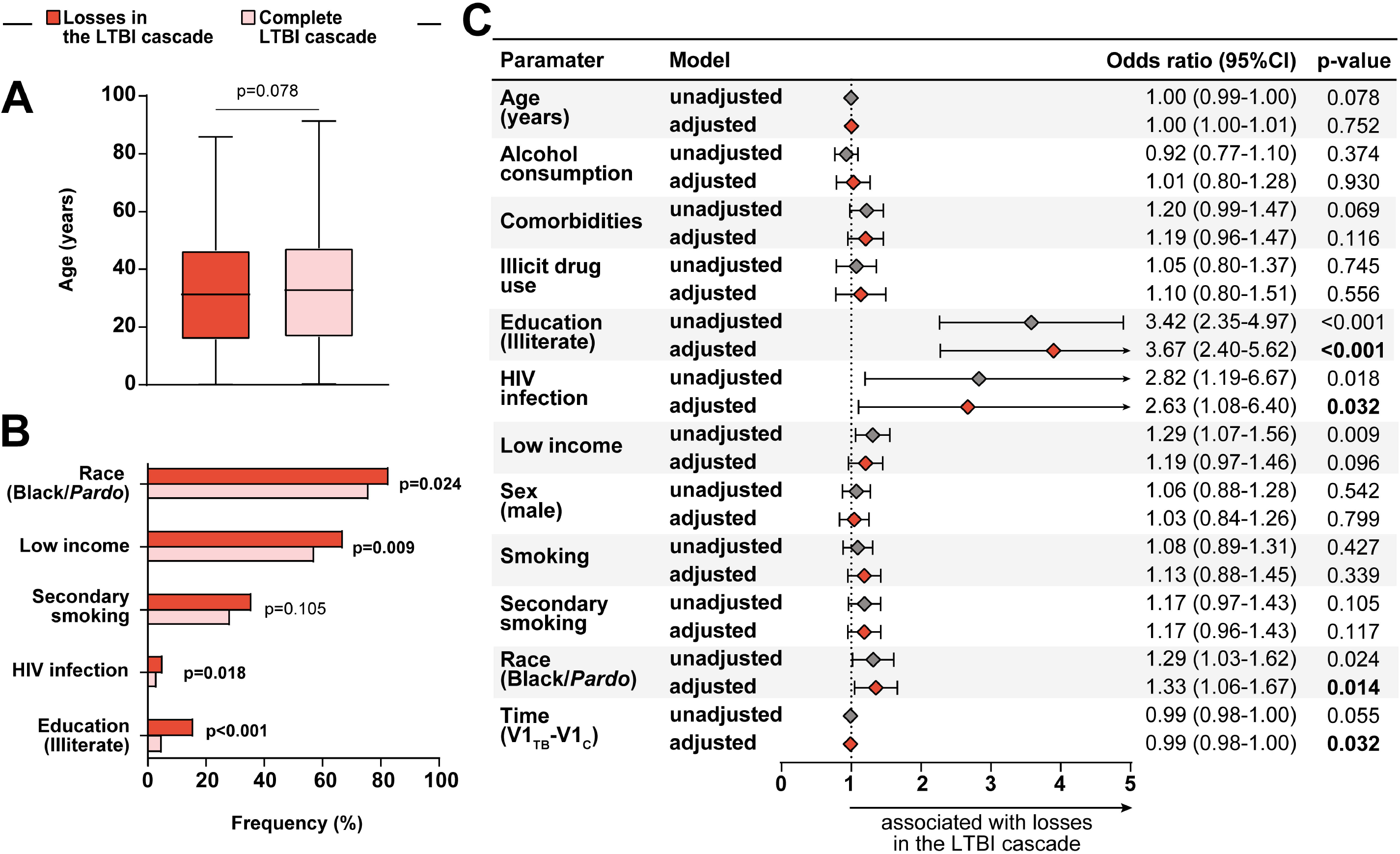
Association between epidemiological and clinical characteristics and losses in the LTBI cascade of care. **(A)** Age distribution among losses in the LTBI cascade of care and those who completed the LTBI cascade. Data were compared using the Mann-Whitney *U* test. **(B)** Frequency of Race/Ethnicity (black and *pardo*), income (see definition below), secondary smoking, comorbidities (see definition below) and education (illiterate) between TB contacts stratified based on losses in the LTBI cascade. Data were compared using Fisher’s exact test. **(C)** Generalized estimating equations analysis to evaluate association between epidemiological and clinical characteristics and losses in the LTBI cascade of care. The study population was stratified according to complete TPT in the LTBI cascade (complete LTBI cascade of care vs. losses in the LTBI cascade of care, see Supplementary Table 1 for detailed univariate comparisons). A multivariable analysis (see Methods for details) was employed with each variable individually (unadjusted) and variables (panels A and C) were included in a multivariable model (adjusted). In all the comparisons, significant p-values are shown in bold-type font. Comorbidities: At least one comorbidity (diabetes, hypertension, cancer, chronic obstructive pulmonary /emphysema, kidney disease, heart disease, liver disease and depression). Low income: without income/equal or less than a minimum wage (reference: more than a minimum wage) Race (Black/*Pardo*) reference: White, Asian, Indian. Abbreviations: 95%CI: 95% confidence interval. Time (V1_TB_-V1_C_): time (in weeks) difference between the visit 1 of the TB case and the visit 1 of the contact. TPT: TB preventive therapy.

### Factors associated with losses at specific stages of the LTBI cascade of care

Considering those participants who were recommended to take TPT but declined (492 of 933; 53%) **(figure 1B)**, they had a lower individual income (p = 0.005), a shorter time between the diagnosis of the TB index case and evaluation of the contact (p=0.027), a higher rate of illiteracy (p<0.001) and a higher rate of secondary smoking (p=0.001). Three of these four variables were independently associated with not initiating TPT: income (aoR:1.36, 95% CI: 1.03-1.80, p =0.032), education (aoR:3.05, 95% CI: 1.92-4.86, p <0.001) and secondary smoking (aoR:1.61, 95% CI: 1.19-2.16, p =0.002) (**Supplementary Figure 2C and Supplementary Table 2**).

Of 454 contacts who initiated TPT, 202 (44%) did not complete TPT **(figure 1A)**. Those who did not complete TPT were younger (p=0.004) and more likely to be black or *pardo* (p=0.012) and lower income (p=0.03) than contacts who completed TPT. Finally, race black or *pardo* (aOR:1.71, 95% CI: 1.01-2.90, p =0.048) was the only factor that remained associated with non-completion of TPT after adjusting for confounding variables (**Supplementary Figure 6 and Supplementary Table 4**).

## DISCUSSION

We found losses in all steps of the cascade, but the most substantial loss occurred before the visit to the clinic, when individuals identified by the TB index cases as contacts did not come for evaluation. This is consistent with findings from previous studies [7]. Two main reasons for this finding have been identified previously: (i) not having interest in being tested and (iii) self-perceived low risk of TB infection [7]. In a study of 1,908 TB contacts, some participants refused the contact investigation because they strongly believed that they did not have TB disease given that they did not have any signs or symptoms [16].

A Brazilian study [17] investigating stigma towards TB in the general population found that 57% of the participants knew that LTBI could occur, and 90% would seek treatment for it, suggesting that knowledge and stigma should not be an important barrier in our population. This factor should be better explored, along with other possible issues such as economic barriers for health care seeking, difficulties pertaining to absences from work and risk perception. Regarding interventions that could reduce losses in the first step of the cascade, one possibility is to increase participation of the Brazilian family health program [18] in searching for contacts. The program includes community agents who are responsible for following a certain number of families and know the community dynamics. This could be a useful tool to better identify and facilitate access of TB contacts with the health care system.

In our study, we found a prevalence of LTBI in close TB contacts of 43%. A similar result was found in a previous study by our group in a different cohort of 1,264 TB contacts and using TST instead of IGRA. The study found a LTBI prevalence of 41.9% and the incidence of active TB of 2/100,000 inhabitants [19]. LTBI was significantly more common in women, black/*pardo*, older individuals, and active and secondhand smokers. Identifying risk factors for LTBI was not within the scope of the current study, but we did identify known risk factors such as tobacco smoking [20], and older age [21]. Interestingly, women were more likely to have LTBI than men, which differs from results reported in other countries [22, 23]. This finding and the fact that black/*pardo* individuals were more likely to be infected than other races merit an additional study, as it could reflect social dynamics in Brazil. The most recent report from the Instituto de *Pesquisa Econômica Aplicada* (IPEA) demonstrated that the number of homes with a female as the referent household member has been increasing over recent decades, but many of these women are homemakers and do not have a formal job [24]. It is possible that women have higher exposure time to sick persons in the household, leading to higher risk of LTBI. The same report identified ethnicity reflecting social vulnerability in Brazil. For instance, black/*pardo* individuals more frequently have lower income and years of education, lower access to sanitation, adequate housing and health care, which could explain our results [25].

In this prospective, multicenter, cohort study we analyzed the cascade of care of LTBI in Brazil and found that HIV infection, illiteracy, low income, and race black or *pardo* were independently associated with greater losses at all stages of the cascade. All of these factors are related to social vulnerability and highlight the relevance of social factors in TB care, previously noted in different studies [26]. These results reinforce the need to consider social context when developing health care public policies.

Modelling studies suggest that diagnosing and treating LTBI in persons at high risk of developing active disease will accelerate TB elimination [4, 27]. However, there can be many challenges in implementing this policy programmatically. Thus, evaluating the cascade of care of LTBI and the factors associated with failures at each step can provide important insights for TB control.

A recently published meta-analysis [28] and literature review analyzing evidence on interventions to reduce loss in LTBI cascade found that completion of the initial assessment (e.g., return for medical visits) is significantly improved by financial and non-financial patient incentives. To a lesser extent, home visits, reminders and health care worker education were also shown to be helpful. Patient incentives and education also improved rates of patient acceptance of LTBI treatment.

Another point worth noting in our study was that less than half of TB contacts who had an indication for TPT accepted and started it. This was lower than what was found in a meta-analysis (72.2%, CI 48-96%) of 6 cohorts from countries at the low- or middle-income level [7]. However, our study included patients who came to the medical visit and refused treatment, as well as patients who never came to get the test results and were not seen by the physician to have the issue approached.

In our study, 54% of TB contacts who initiated TPT completed it. A similar proportion (56%) was observed in a cohort of 336 TB contacts in a primary health care service in São Paulo state, Brazil and in studies in low or middle-income countries (about 50%) [7, 9]. We found that older age and lower income were associated with abandonment of TPT. Barriers to treatment completion demonstrated before are diverse and vary according to location. Common barriers include side-effects to drugs, long treatment duration, issues related to the health-system and individual concerns, such as substance use [7,9]. Financial barriers and low level of knowledge about the cost-benefit of treatment have also been associated with poor treatment adherence in LTBI patients [29].

Our study had several limitations. Because the contacts were enrolled in a study conducted in referral centers, and under research conditions, the results may not be generalizable to all close contacts of TB index cases. In addition, we did not capture the proportion of physicians who did or did not recommend TPT according to Brazilian Guidelines.

With the above limitations noted, the results presented here illuminate substantial losses to follow up among a population at high risk of developing active TB disease. Mitigating losses in the LTBI care cascade is an important step for the control and eradication of TB.

## Data Availability

All data used is available through contact with the corresponding author.

## Notes

## Acknowledgments

The authors thank the study participants. We also thank the teams of clinical and laboratory platforms of RePORT Brazil. A special thanks to Elze Leite (FIOCRUZ, Salvador, Brazil), Eduardo Gama (FIOCRUZ, Rio de Janeiro, Brazil), Elcimar Junior (FMT-HVD, Manaus, Brazil) and Hilary Vansell (VUMC, Nashville, USA), for administrative and logistical support.

## Disclaimer

The funders of the study had no role in study design, data analysis, data interpretation, or writing of the report. All authors had access to all the data in the study and had final responsibility for the decision to submit for publication.

