## Supplemental material for "Determinants of losses in the latent tuberculosis infection cascade of care in Brazil: a prospective multicenter cohort study"

**Supplementary Material**

**Content:**

1. **Supplementary Table 1**
2. **Supplementary Table 2**
3. **Supplementary Table 3**
4. **Supplementary Table 4**
5. **Supplementary Figure 1**
6. **Supplementary Figure 2**
7. **Supplementary Figure 3**
8. **Supplementary Figure 4**

**Supplementary Tables**

**Supplementary Table 1. Characteristics of contacts according to losses and completeness of LTBI cascade of care**

| **Characteristics** | **Lost in the cascade**  **(n=895)*** | **Completed the cascade**  **(n=1001)** | **p-value** |
| --- | --- | --- | --- |
| **Female– no. (%)** | 538 (60) | 589 (59) | 0.542 |
| **Age – median (IQR)** | 31 (16-47) | 33 (17-47) | 0.078 |
| **Race/Ethnicity – no. (%)** |  |  | 0.024 |
| Black/ Pardo | 741 (83) | 761 (76) |  |
| Others | 153 (17) | 240 (24) |  |
| **Income – no. (%)** |  |  | 0.009 |
| More than a minimum wage | 285 (33) | 412 (43) |  |
| Equal or less than a minimum wage | 386 (44) | 365 (38) |  |
| Without income | 197 (23) | 189 (19) |  |
| **BCG scar – no. (%)** | 802 (90) | 893 (89) | 0.820 |
| **Time (V1_TB_-V1_C_)** | 33 (8-82) | 41 (14-102) | 0.055 |
| **HIV infection – no. (%)** | 31 (4) | 15 (2) | 0.018 |
| **Antiretroviral therapy – no. (%)** | 23 (74) | 13 (87) | 0.460 |
| **Education– no. (%)** |  |  | <0.001 |
| Literate | 753 (84) | 950 (95) |  |
| Illiterate | 141 (16) | 50 (5) |  |
| **Smoking – no. (%)** | 247 (28) | 254 (25) | 0.427 |
| **Secondary smoking – no. (%)** | 319 (36) | 284 (28) | 0.105 |
| **Alcohol consumption – no. (%)** | 471 (53) | 545 (54) | 0.433 |
| **Alcohol consumption (years)-median (IQR)** | 12 (4-22) | 11 (3-20) | 0.337 |
| **CAGE score of 2 and above** ^a^ **– no. (%)** | 82 (26) | 96 (29) | 0.333 |
| **Illicit drug use – no. (%)** | 91 (10) | 97 (10) | 0.758 |
| **Comorbidities^b^– no. (%)** | 241 (27) | 204 (20) | 0.069 |
| Diabetes | 43 (5) | 52 (5) | 0.752 |
| Hypertension | 132 (15) | 101 (10) | 0.003 |

**Table note:** Data represent no. (%), except for age time, which is presented as median and interquartile range (IQR) Continuous variables were compared using the Mann-Whitney *U* test and categorical variables were using the Fisher's exact test (2x2) or Pearson’s chi-square test. Time (V1_TB_-V1_C_): time (in weeks) difference between the visit 1 of the TB case and the visit 1 of the contact.

**^*^**Five contacts are ongoing in the TPT

^a^Alcohol abuse was defined as a CAGE score ≥ 2 points *(Ewing, J.A. 1984. Detecting alcoholism: The CAGE questionnaire. JAMA).*

^b^ Comorbidities: At least one comorbidity (diabetes, hypertension, cancer, chronic obstructive pulmonary /emphysema, kidney disease, heart disease, liver disease and depression)

Abbreviations: TB: tuberculosis. LTBI: latent tuberculosis infection. TPT: TB preventive therapy

**Supplementary Table 2. Characteristics of contacts according to Initiation of recommended TPT**

| **Characteristics** | **No initiation of recommended TPT**  **(n=492)** | **Initiation of recommended TPT**  **(n=454*)** | **p-value** |
| --- | --- | --- | --- |
| **Female– no. (%)** | 299 (61) | 272 (60) |  |
| **Age – median (IQR)** | 32.5 (11-48) | 32.6 (15-48) | 0.902 |
| **Race/Ethnicity – no. (%)** |  |  | 0.247 |
| Black/ Pardo | 408 (83) | 362 (80) |  |
| Others | 83 (17) | 92 (20) |  |
| **Income – no. (%)** |  |  | 0.005 |
| More than a minimum wage | 150 (31) | 171 (39) |  |
| Equal or less than a minimum wage | 236 (49) | 175 (40) |  |
| Without income | 94 (20) | 92 (21) |  |
| **BCG scar – no. (%)** | 444 (90) | 400 (88) | 0.296 |
| **Time (V1_TB_-V1_C_)** | 24 (7-71) | 45 (15-102) | 0.027 |
| **HIV infection – no. (%)** | 19 (4) | 26 (6) | 0.555 |
| **Antiretroviral therapy – no. (%)** | 12 (63) | 23 (89) | 0.070 |
| **Education– no. (%)** |  |  | <0.001 |
| Literate | 379 (77) | 404 (89) |  |
| Illiterate | 113 (23) | 50 (11) |  |
| **Smoking – no. (%)** | 131 (27) | 134 (30) | 0.911 |
| **Secondary smoking – no. (%)** | 190 (39) | 134 (30) | 0.001 |
| **Alcohol consumption – no. (%)** | 233 (47) | 233 (51) | 0.241 |
| **Alcohol consumption (years)-median (IQR)** | 14 (5-21) | 13 (4-23) | 0.918 |
| **CAGE score of 2 and above** ^a^ **– no. (%)** | 31 (20) | 40 (29) | 0.133 |
| **Illicit drug use – no. (%)** | 37 (8) | 54 (12) | 0.128 |
| **Comorbidities ^b^– no. (%)** | 128 (26) | 102 (23) | 0.295 |
| Diabetes | 23 (5) | 20 (4) | 0.880 |
| Hypertension | 65 (13) | 64 (14) | 0.705 |

**Table note:** Data represent no. (%), except for age and time, which is presented as median and interquartile range (IQR) Continuous variables were compared using the Mann-Whitney *U* test and categorical variables were using the Fisher's exact test (2x2) or Pearson’s chi-square test. Time (V1_TB_-V1_C_): time (in weeks) difference between the visit 1 of the TB case and the visit 1 of the contact.

* Among those to whom TPT was recommended (n=933)

^a^ Alcohol abuse was defined as a CAGE score ≥ 2 points as described in Methods*.*

^b^ Comorbidities: At least one comorbidity (diabetes, hypertension, cancer, chronic obstructive pulmonary /emphysema, kidney disease, heart disease, liver disease and depression)

Abbreviations: TB: tuberculosis. TPT: TB preventive therapy

**Supplementary Table 3. Characteristics of contacts according to complete TB preventive therapy**

| **Characteristics** | **Incomplete TPT**  **(n=202*)** | **Complete**  **TPT**  **(n=247)** | **p-value** |
| --- | --- | --- | --- |
| **Female– no. (%)** | 116 (57) | 153 (62) | 0.470 |
| **Age – median (IQR)** | 28.2 (15-43) | 37.5 (15-53) | 0.004 |
| **Race/Ethnicity – no. (%)** |  |  | 0.012 |
| Black/ Pardo | 172 (85) | 185 (75) |  |
| Others | 30 (15) | 62 (25) |  |
| **Income – no. (%)** |  |  | 0.030 |
| More than a minimum wage | 66 (34) | 105 (44) |  |
| Equal or less than a minimum wage | 82 (42) | 92 (39) |  |
| Without income | 47 (24) | 41 (17) |  |
| **BCG scar – no. (%)** | 176 (87) | 219 (89) | 0.663 |
| **Time (V1_TB_-V1_C_)** | 45 (14-92) | 47 (17-106) | 0.217 |
| **HIV infection – no. (%)** | 12 (6) | 14 (6) | 0.361 |
| **Antiretroviral therapy – no. (%)** | 11 (92) | 12 (86) | 1.000 |
| **Education– no. (%)** |  |  | 0.982 |
| Literate | 176 (87) | 224 (91) |  |
| Illiterate | 26 (13) | 23 (9) |  |
| **Smoking – no. (%)** | 55 (27) | 77 (31) | 0.195 |
| **Secondary smoking – no. (%)** | 63 (31) | 69 (28) | 0.641 |
| **Alcohol consumption – no. (%)** | 111 (55) | 119 (48) | 0.156 |
| **Alcohol consumption (years)-median (IQR)** | 12 (3-24) | 14 (4-23) | 0.581 |
| **CAGE score of 2 and above** ^a^ **– no. (%)** | 26 (34) | 12 (19) | 0.058 |
| **Illicit drug use – no. (%)** | 27 (13) | 25 (10) | 0.876 |
| **Comorbidities^b^– no. (%)** | 55 (27) | 47 (19) | 0.119 |
| Diabetes | 6 (3) | 14 (6) | 0.250 |
| Hypertension | 35 (17) | 29 (12) | 0.104 |

**Table note:** Data represent no. (%), except for age and time, which is presented as median and interquartile range (IQR) Continuous variables were compared using the Mann-Whitney *U* test and categorical variables were using the Fisher's exact test (2x2) or Pearson’s chi-square test. Time (V1_TB_-V1_C_): time (in weeks) difference between the visit 1 of the TB case and the visit 1 of the contact.

*4 no recommended

^a^ Alcohol abuse was defined as a CAGE score ≥ 2 points as described in Methods

^b^ Comorbidities: At least one comorbidity (diabetes, hypertension, cancer, chronic obstructive pulmonary /emphysema, kidney disease, heart disease, liver disease and depression)

Abbreviations: TB: tuberculosis. LTBI: latent tuberculosis infection. TPT: TB preventive therapy

**Supplementary Table 4. Characteristics of contacts according to 2^nd^ IGRA performed**

| **Characteristics** | **2^nd^ IGRA not performed (n=242)** | **2^nd^ IGRA performed**  **(n= 967)** | **p-value** |
| --- | --- | --- | --- |
| **Female– no. (%)** | 142 (59) | 548 (57) | 0.836 |
| **Age – median (IQR)** | 27.1 (16-44.) | 30.3 (15-45) | 0.412 |
| **Race/Ethnicity – no. (%)** |  |  | 0.883 |
| Black/ Pardo | 193 (80) | 749 (78) |  |
| Others | 48 (20) | 218 (23) |  |
| **Income – no. (%)** |  |  | 0.048 |
| More than a minimum wage | 81 (35) | 392 (42) |  |
| Equal or less than a minimum wage | 83 (36) | 362 (39) |  |
| Without income | 69 (29) | 182 (19) |  |
| **BCG scar – no. (%)** | 221 (91) | 865 (90) | 0.475 |
| **Time (V1_TB_-V1_C_)** | 36 (15-96) | 40 (13-99) | 0.955 |
| **HIV infection – no. (%)** | 7 (3) | 28 (3) | 0.883 |
| **Antiretroviral therapy – no. (%)** | 5 (71) | 24 (86) | 0.576 |
| **Education– no. (%)** |  |  | 0.632 |
| Literate | 211 (88) | 860 (89) |  |
| Illiterate | 30 (12) | 106 (11) |  |
| **Smoking – no. (%)** | 66 (27) | 227 (24) | 0.301 |
| **Secondary smoking – no. (%)** | 76 (32) | 284 (30) | 0.872 |
| **Alcohol consumption – no. (%)** | 133 (55 | 510 (53) | 0.565 |
| **Alcohol consumption (years)-median (IQR)** | 10 (2-18) | 10 (3-18) | 0.644 |
| **CAGE score of 2 and above** ^a^ **– no. (%)** | 26 (27) | 101 (32) | 0.448 |
| **Illicit drug use – no. (%)** | 29 (12) | 91 (9) | 0.231 |
| **Comorbidities^b^– no. (%)** | 73 (30) | 196 (20) | 0.023 |
| Diabetes | 19 (8) | 43 (4) | 0.049 |
| Hypertension | 40 (17) | 93 (10) | 0.004 |

**Table note:** Data represent no. (%), except for age and time, which is presented as median and interquartile range (IQR). Continuous variables were compared using the Mann-Whitney *U* test and categorical variables were using the Fisher's exact test (2x2) or Pearson’s chi-square test. Time (V1_TB_-V1_C_): time (in weeks) difference between the visit 1 of the TB case and the visit 1 of the contact.

^a^ Alcohol abuse was defined as a CAGE score ≥ 2 points as described in Methods.

^b^ Comorbidities: At least one comorbidity (diabetes, hypertension, cancer, chronic obstructive pulmonary /emphysema, kidney disease, heart disease, liver disease and depression)

Abbreviations: TB: tuberculosis; IGRA: interferon-gamma release assay.

**Supplementary Figures**

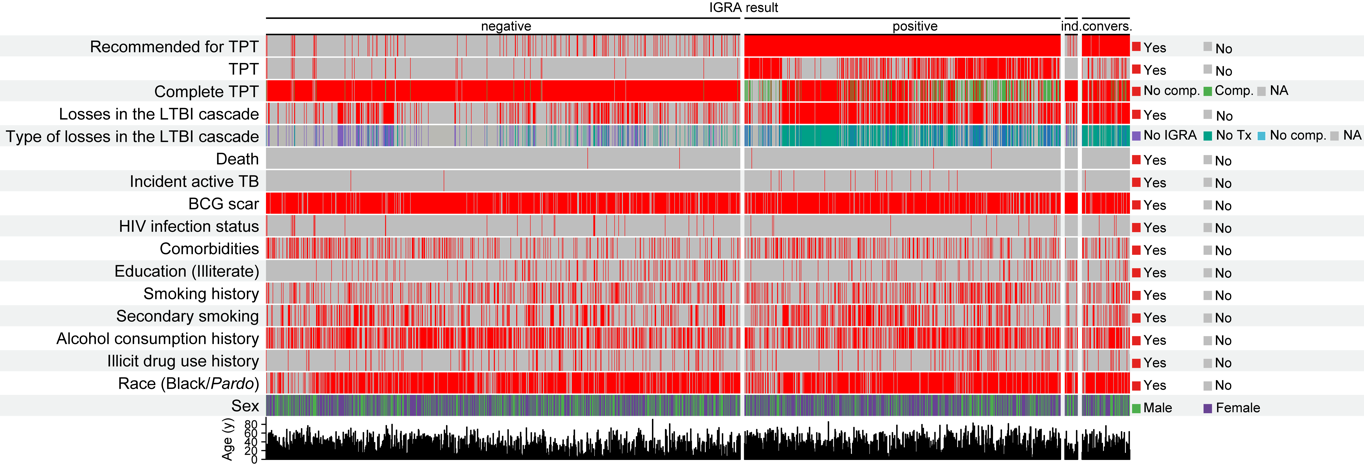

**Supplementary Figure 1. Characteristics of study participants**

Color map based on TB contacts grouped according to final IGRA results shows the overall characteristics of the study participants. A hierarchical clustering analysis (Ward’s method) was employed to group individuals based on the overall profile of each study participant which each IGRA subgroup.

Alcohol abuse was defined as a CAGE score ≥ 2 points. Comorbidities: At least one comorbidity (diabetes, hypertension, cancer, chronic obstructive pulmonary /emphysema, kidney disease, heart disease, liver disease and depression

Abbreviations: IGRA: interferon-gamma release assay, Ind: Indeterminate, Convers: conversion, No IGRA 2: Did not perform 2nd IGRA, LTBI: latent tuberculosis infection, TB: tuberculosis. TPT: TB preventive therapy.

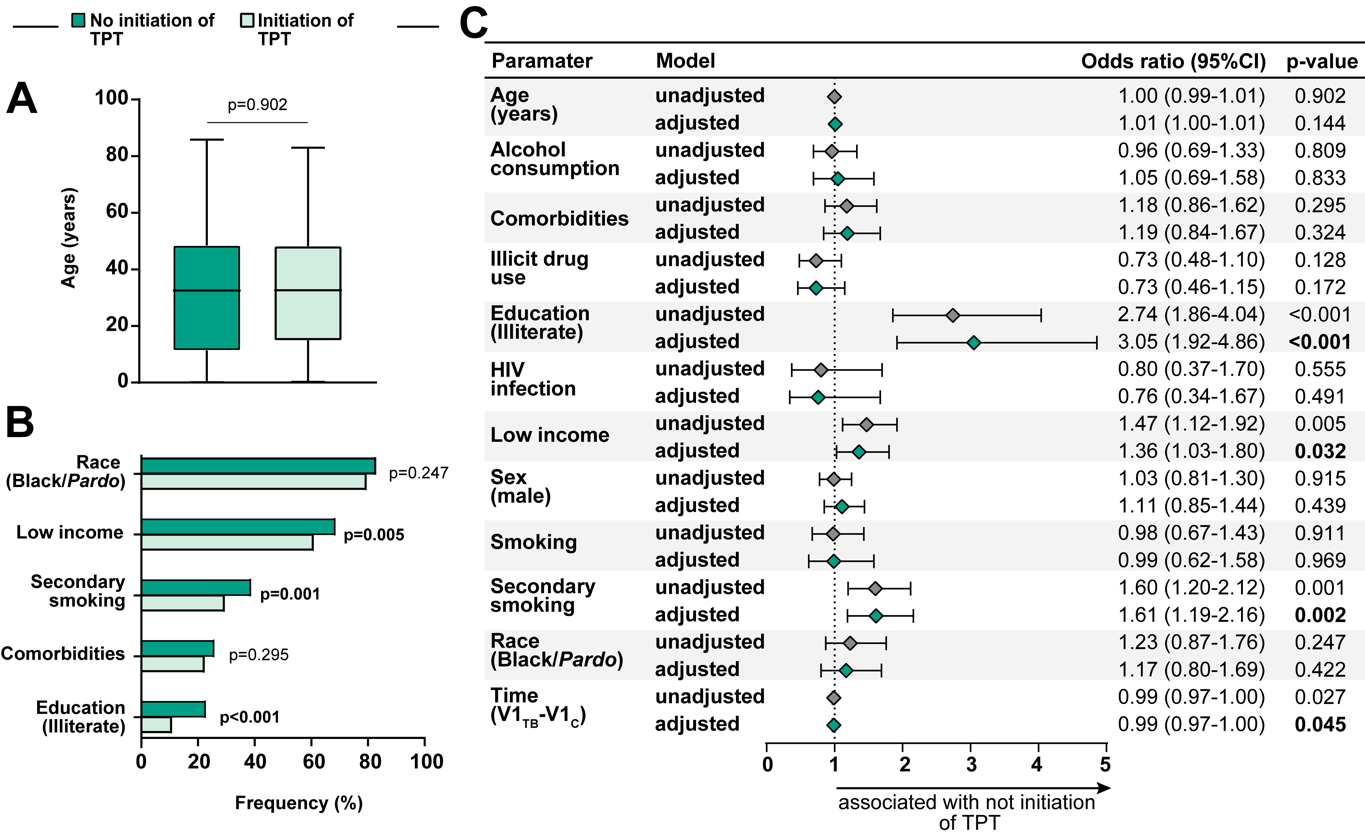

**Supplementary Figure 2. Association between epidemiological and clinical characteristics and initiation of TPT**

**(A)** Age distribution among TB contacts who did not initiate TPT and those who initiated the TPT. Data were compared using the Mann-Whitney *U* test. **(B)** Frequency of Race/Ethnicity (black and *pardo*), income (see definition below), secondary smoking, comorbidities (see definition below) and education (illiterate) between TB contacts stratified based on initiation of TPT**.** Data were compared using Fisher's exact test. **(C)** Generalized estimating equations analysis to evaluate association between epidemiological and clinical characteristics and not initiation of TPT**.** The study population was stratified according to initiation of TPT (not initiation of TPT and initiation of the TPT, see Supplementary Table 2 for detailed univariate comparisons). A multivariable analysis (see Methods for details) was employed with each variable individually (unadjusted) and variables (Panels A and **C)** were included in a multivariable model (adjusted). In all the comparisons, significant p-values are shown in bold-type font.

Comorbidities: At least one comorbidity (diabetes, hypertension, cancer, chronic obstructive pulmonary /emphysema, kidney disease, heart disease, liver disease and depression).

Low income: without income/equal or less than a minimum wage (reference: more than a minimum wage)

Race (Black/*Pardo*) reference: White, Asian, Indian.

Abbreviations: 95%CI: 95% confidence interval. Time (V1TB-V1C): time (in weeks) difference between the visit 1 of the TB case and the visit 1 of the contact. TPT: TB preventive therapy.

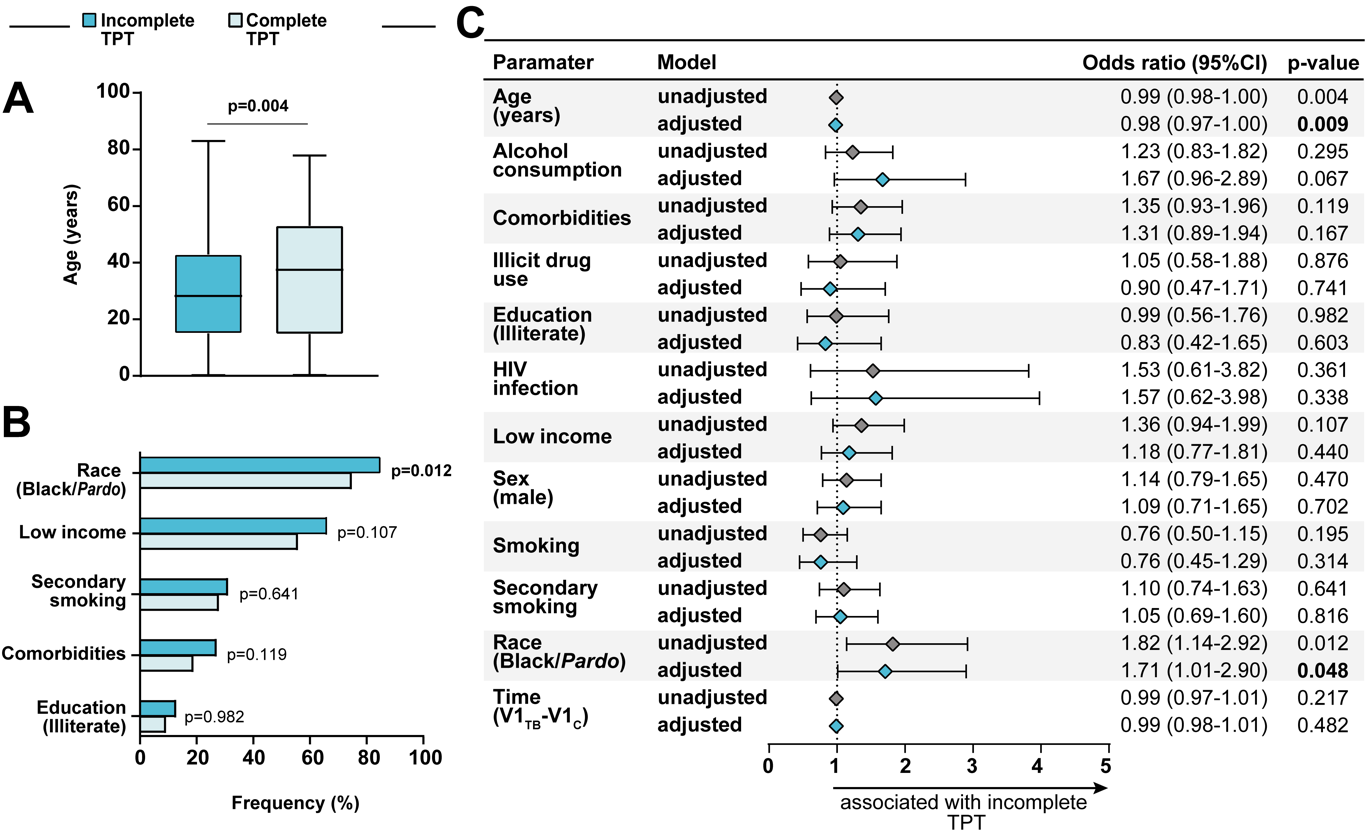

**Supplementary Figure 3. Association between epidemiological and clinical characteristics and completeness of the TB preventive therapy**

**(A)** Age distribution among those who did not complete the TPT and who complete the TPT. Data were compared using the Mann-Whitney *U* test. **(B)** Frequency of Race/Ethnicity (black and *pardo*), income (see definition below), secondary smoking, comorbidities (see definition below) and education (illiterate) between TB contacts stratified based completeness of the TPT (complete vs. incomplete)**.** Data were compared using Fisher's exact test. **(C)** Generalized estimating equations analysis to evaluate association between epidemiological and clinical characteristics and who did not complete the TPT and who complete the TPT. The study population was stratified according to complete TPT in the LTBI cascade (incomplete TPT vs. complete cascade of care, see Supplementary Table 3 for detailed univariate comparisons). A multivariable analysis (see Methods for details) was employed with each variable individually (unadjusted) and variables (panels A and **C)** were included in a multivariable model (adjusted). In all the comparisons, significant p-values are shown in bold-type font.

Comorbidities: At least one comorbidity (diabetes, hypertension, cancer, chronic obstructive pulmonary /emphysema, kidney disease, heart disease, liver disease and depression).

Low income: without income/equal or less than a minimum wage (reference: more than a minimum wage)

Race (Black/*Pardo*) reference: White, Asian, Indian.

Abbreviations: 95%CI: 95% confidence interval. Time (V1_TB_-V1_C_): time (in weeks) difference between the visit 1 of the TB case and the visit 1 of the contact. TPT: TB preventive therapy.

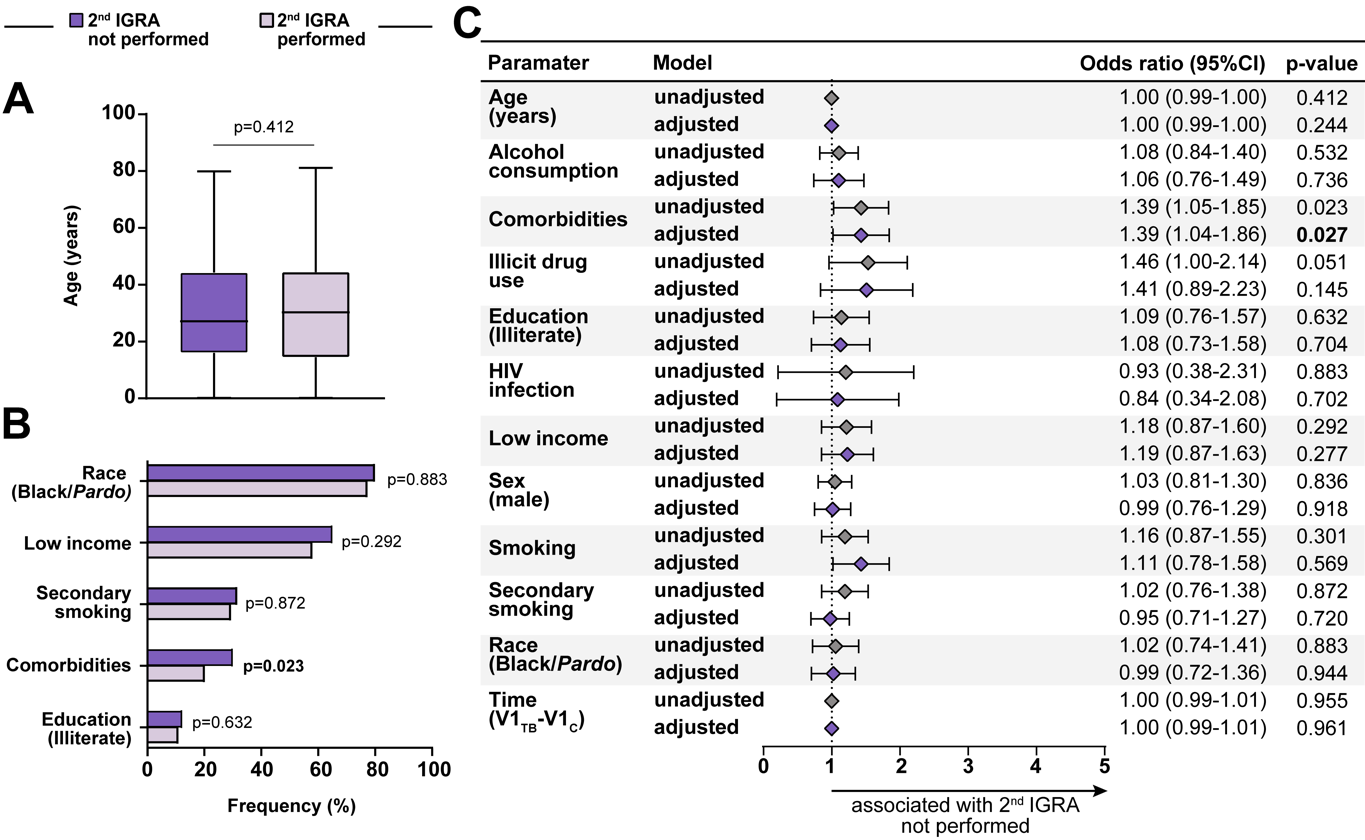

**Supplementary Figure 4. Association between epidemiological and clinical characteristics and performing the 2^nd^ IGRA test at month 6 when indicated. (A)** Age distribution among those who performed the 2^nd^ IGRA and those not performed the 2^nd^ IGRA. Data were compared using the Mann-Whitney *U* test. **(B)** Frequency of Race/Ethnicity (black and *pardo*), income (see definition below), secondary smoking, comorbidities (see definition below) and education (illiterate) between TB contacts stratified based on performing the 2^nd^ IGRA**.** Data were compared using the Fisher’s exact test. **(C)** Generalized estimating equations analysis to evaluate association between epidemiological and clinical characteristics and losses in the LTBI cascade of care**.** The study population was stratified according to perform 2^nd^ IGRA (2^nd^ IGRA not performed and 2^nd^ IGRA performed, see Supplementary Table 3 for detailed univariate comparisons). A multivariable analysis (see Methods for details) was employed with each variable individually (unadjusted) and variables (panels A and C) were included in a multivariable model (adjusted). In all the comparisons, significant p-values are shown in bold-type font.

Comorbidities: At least one comorbidity (diabetes, hypertension, cancer, chronic obstructive pulmonary /emphysema, kidney disease, heart disease, liver disease and depression).

Low income: without income/equal or less than a minimum wage (reference: more than a minimum wage)

Race (Black/*Pardo*) reference: White, Asian, Indian.

Abbreviations: 95%CI: 95% confidence interval. Time (V1TB-V1C): time (in weeks) difference between the visit 1 of the TB case and the visit 1 of the contact.
